## Supplementary Figures and Text for "Genetic influences on circulating retinol and its relationship to human health"

**Supplementary Materials**


Supplementary Figures 1-12 2

Supplementary Text 17

### SUPPLEMENTARY FIGURES

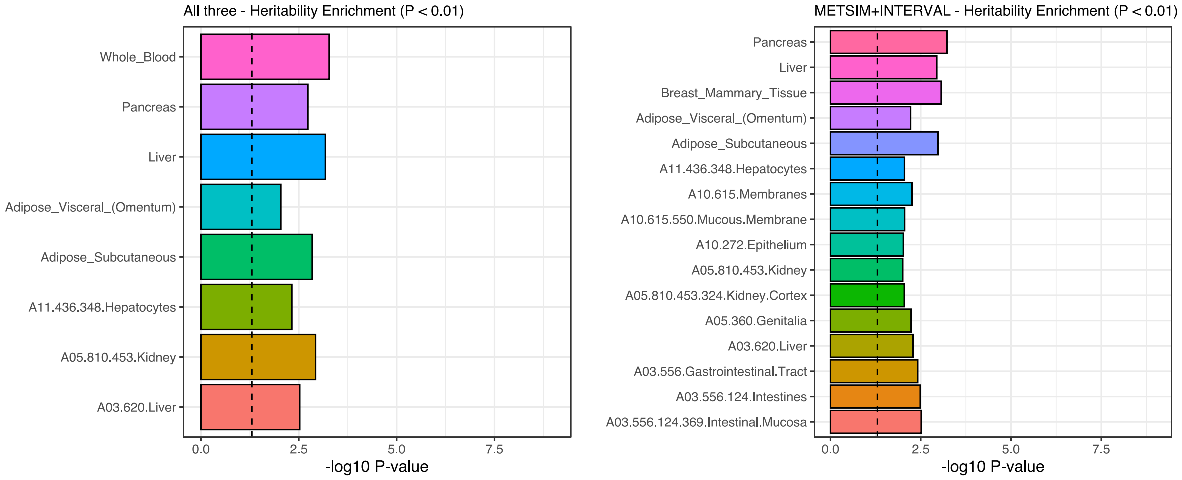


**Supplementary Figure 1. Partitioned SNP heritability estimates amongst diverse tissues and cell-types for circulating retinol.** The barplot denotes tissues or cell types which there was at least some nominal evidence ($P < 0.01$) of significant enrichment of heritability. The left-hand plot visualises results from analysis of the larger GWAS (METSIM+INTERVAL+ATBC+PLCO), whilst the METSIM+INTERVAL meta-analysis is on the righthand side.

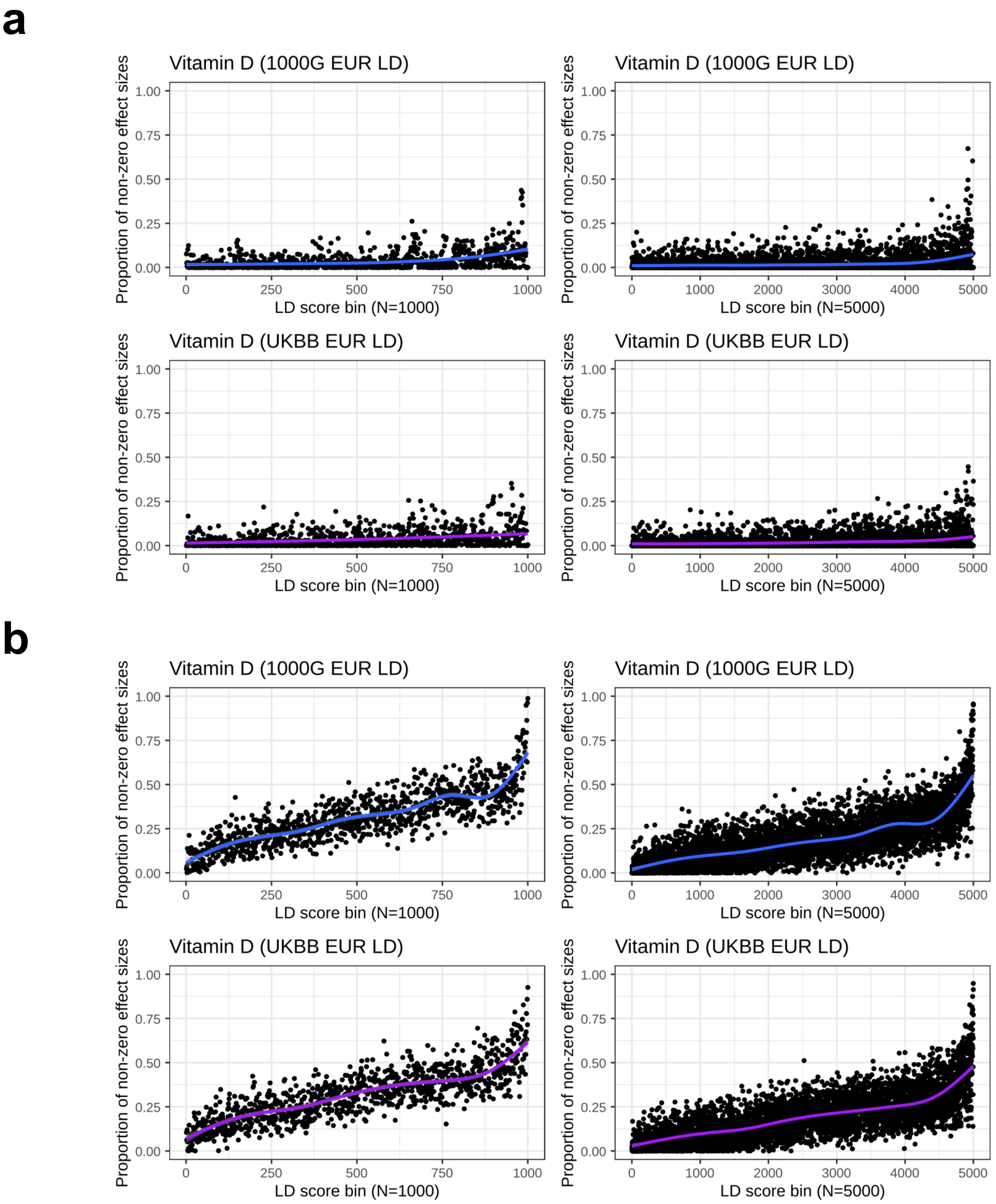


**Supplementary Figure 2. Empirical Bayes' estimation of non-null effects on vitamin D genome-wide, stratified by bins of ascendingly sorted LD score by magnitude.** Analysis performed on two vitamin D GWAS of comparable sample size to the retinol GWAS (**a**) and a much larger GWAS (**b**). The GWAS in (**a**) is from the GRASP consortium with a sample size up to 42,274, whilst a UK Biobank GWAS of vitamin D is analysed in (**b**) with a sample size of up to 417,580. For (**a**) and (**b**), the LD score bins were different for each panel – 1000 bins, 1000 genomes European LD scores (top left); 5000 bins, 1000 genomes European LD scores (top right); 1000 bins, UKBB white British LD scores (bottom left); 5000 bins, white British LD scores (bottom right). Each point represents the proportion of non-null effect sizes for that bin, with the trendline estimated using a generalised additive model for the relationship between the LD score bin and the proportion of non-null effects.

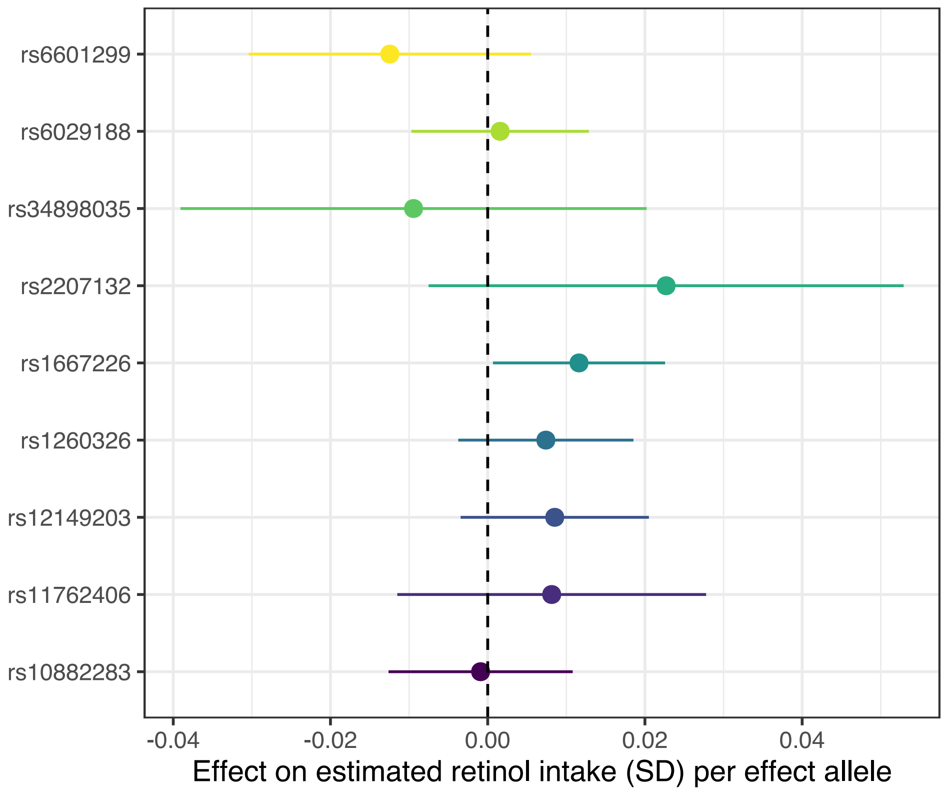


**Supplementary Figure 3. The effect of circulating retinol associated lead SNPs on retinol intake.** Forest plot denotes the point-estimate, with 95% confidence intervals (error bars), of each lead SNP on a GWAS of estimated retinol intake as derived from the responses to a 24 hour dietary recall by UK Biobank participants.

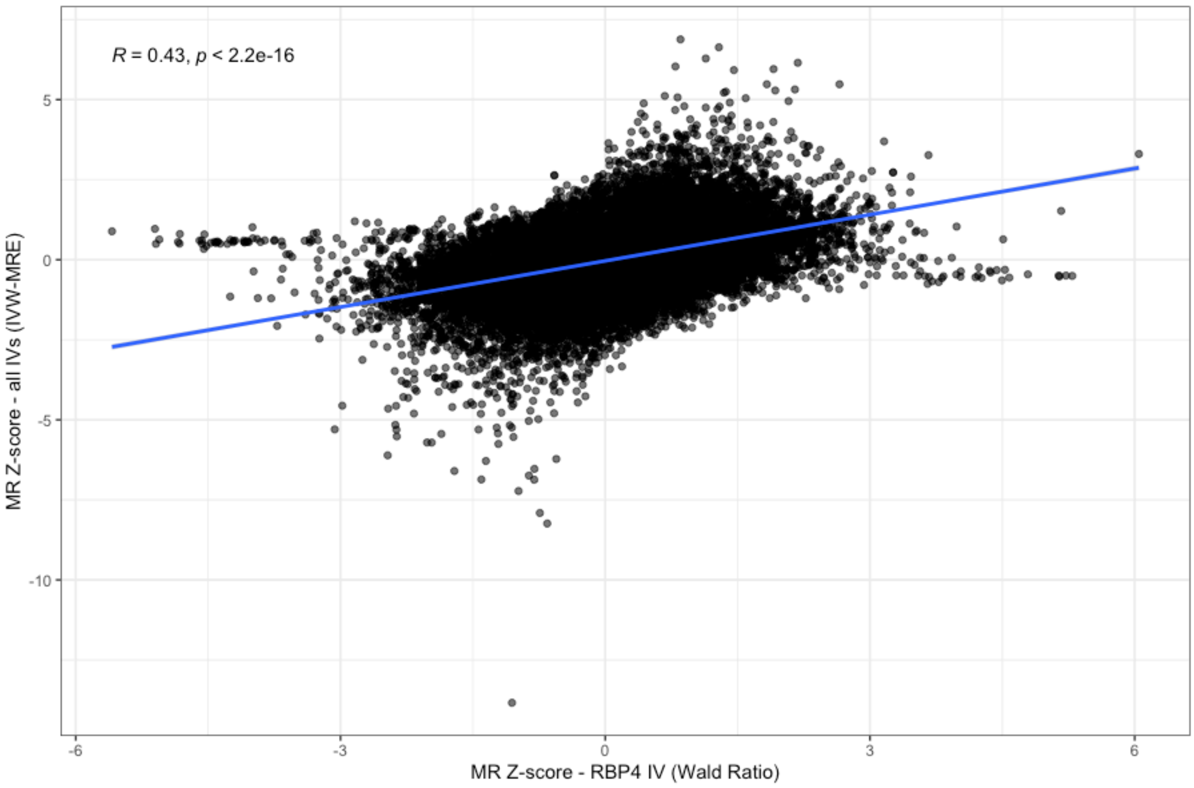


**Supplementary Figure 4. Correlation between normalised effect sizes of phenome-wide Mendelian randomisation (MR Z-score) using a single IV in *RBP4* versus all independent genome-wide significant SNPs as IVs**. Each point denotes an MR Z-score for a retinol-outcome estimate, with a linear trend line plotted and a Pearson's correlation coefficient denoted on the plot.

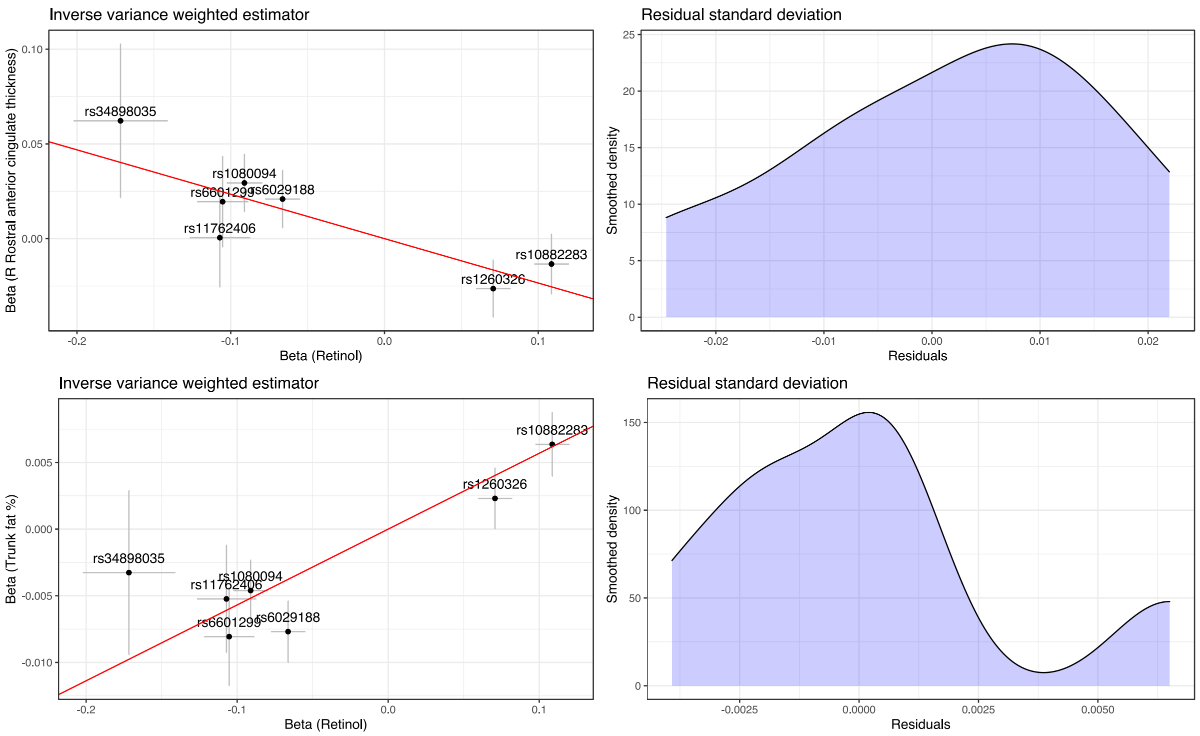


**Supplementary Figure 5. Exploring the effect of residual standard error on the IVW-multiplicative random effects (MRE) estimates relative to that of the IVW fixed effects (FE) estimates**. The top right plot indicates the individual IV exposure-outcome relationships and IVW slope for an outcome trait (right rostral anterior caudate thickness) where the standard errors differ by a noticeable amount between the IVW-MRE and IVW-FE – however, both are still statistically significant (Supplementary Table 20). The left-plot denotes the smoothed density of residuals, with a relatively small spread of values. In contrast, the second row of panels denotes an outcome where the IVW-MRE and IVW-FE standard errors are very similar (trunk fat percentage). This is because the residual standard deviation is larger and closer to one.

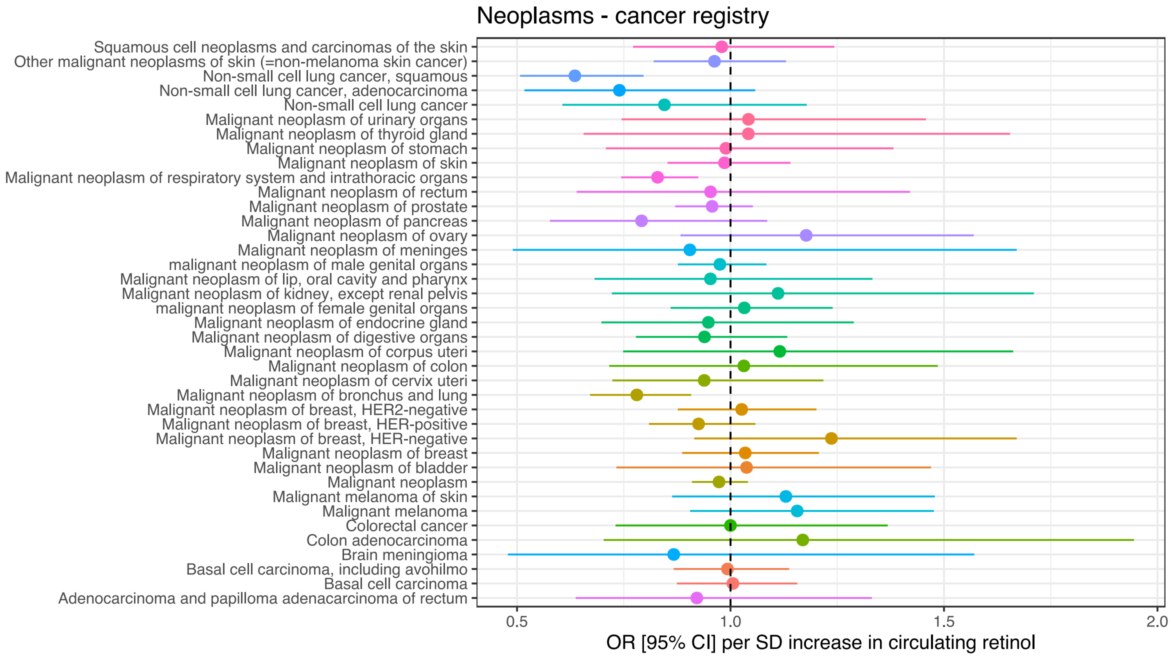


**Supplementary Figure 6. Mendelian randomisation causal estimates (IVW-MRE) of circulating retinol on cancer registry outcomes in FinnGen release 8**. Forest plot denotes the causal estimate (odds ratio, with 95% CI) on each cancer outcome.

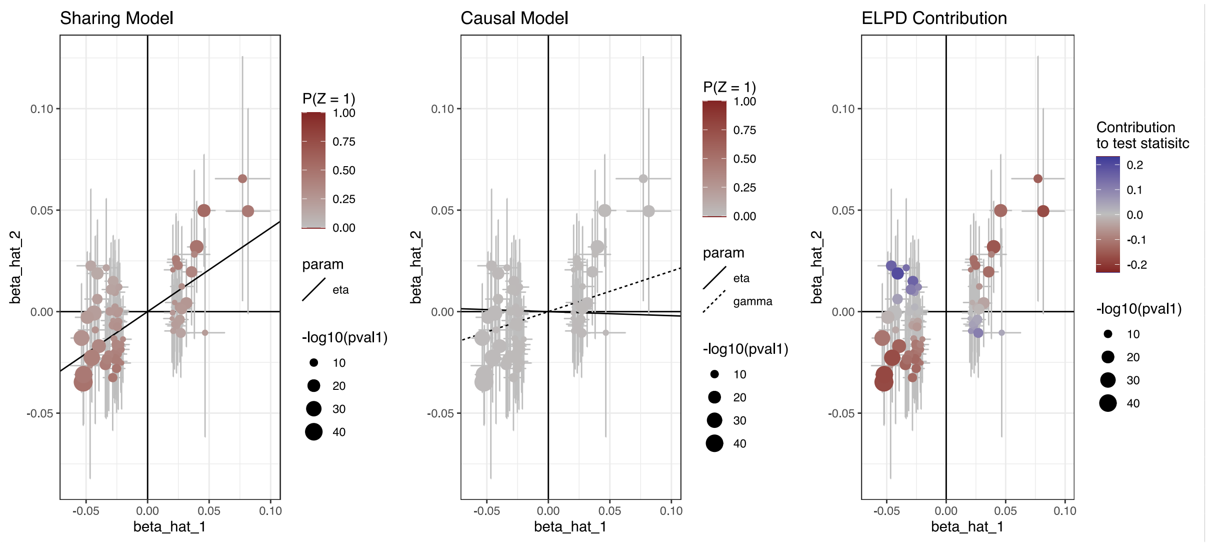


**Supplementary Figure 7. Results from comparing competing models in the CAUSE framework for the effect of serum creatinine on circulating retinol.** The sharing model (eta) is shown on the left-most panel, whilst the causal model (eta and gamma) is in the middle. The contribution of each variant to the ELPD model comparison is denoted in the right panel.

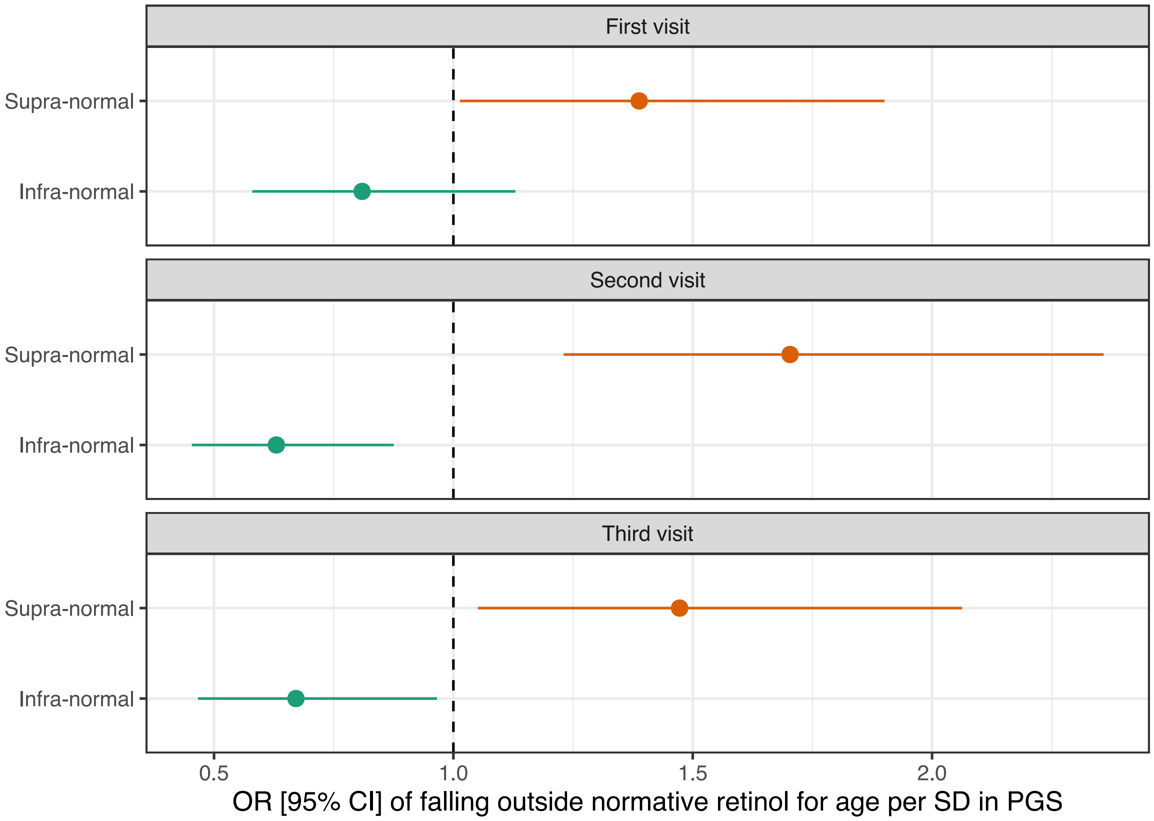


**Supplementary Figure 8. The association between retinol PGS and falling outside of the normative range of measured circulating retinol for a given age**. Forest plot denotes the odds ratios and 95% confidence intervals (error bars) from binomial logistic regression models that tested the association between retinol PGS with normative deviations for age. Specifically, this was from a GAMLSS normative model applied only to unrelated individuals (half of twins). Supra-informal individuals have circulating retinol above the 95$^{th}$ percentile, whilst infra-normal relates to values below the 5$^{th}$ percentile. The dotted line denotes the null odds ratio of one.

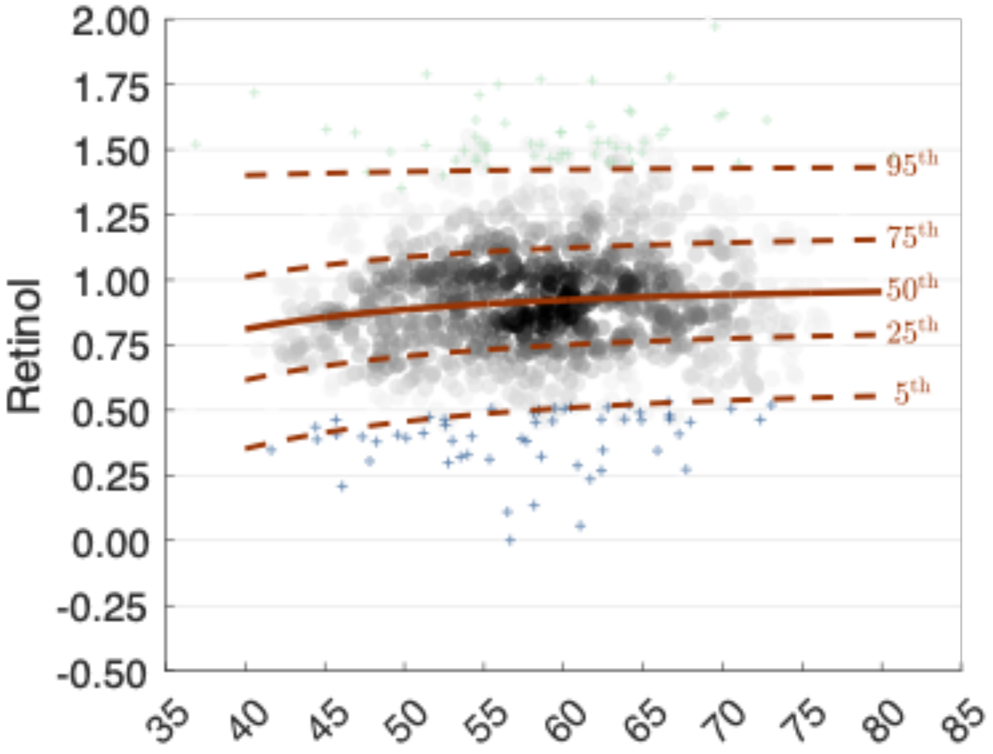


**Supplementary Figure 9. Example of normative model of measured circulating retinol with age**. Data derived from second visit amongst unrelated participants only (half of the twins). The plot denotes normative centile ranges, with quantiles inferred using a GAMLSS model as described in text. Individuals classified as infra-normal as coloured blue, whilst supra-normal individuals are coloured light green.

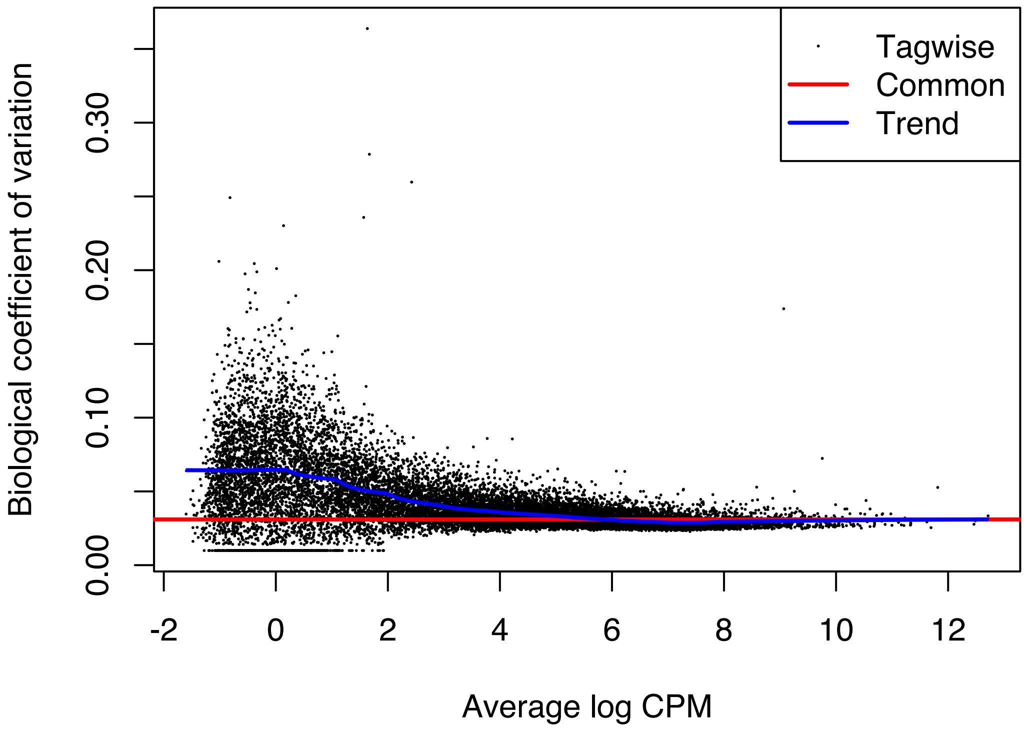


**Supplementary Figure 10. Genewise plot of the biological coefficient of variation (BCV) versus transcript abundance log counts per million (CPM).** Data derived from FOXP2 overexpression experiment, as described in the main text. Common, trend, and tagwise dispersion plotted.

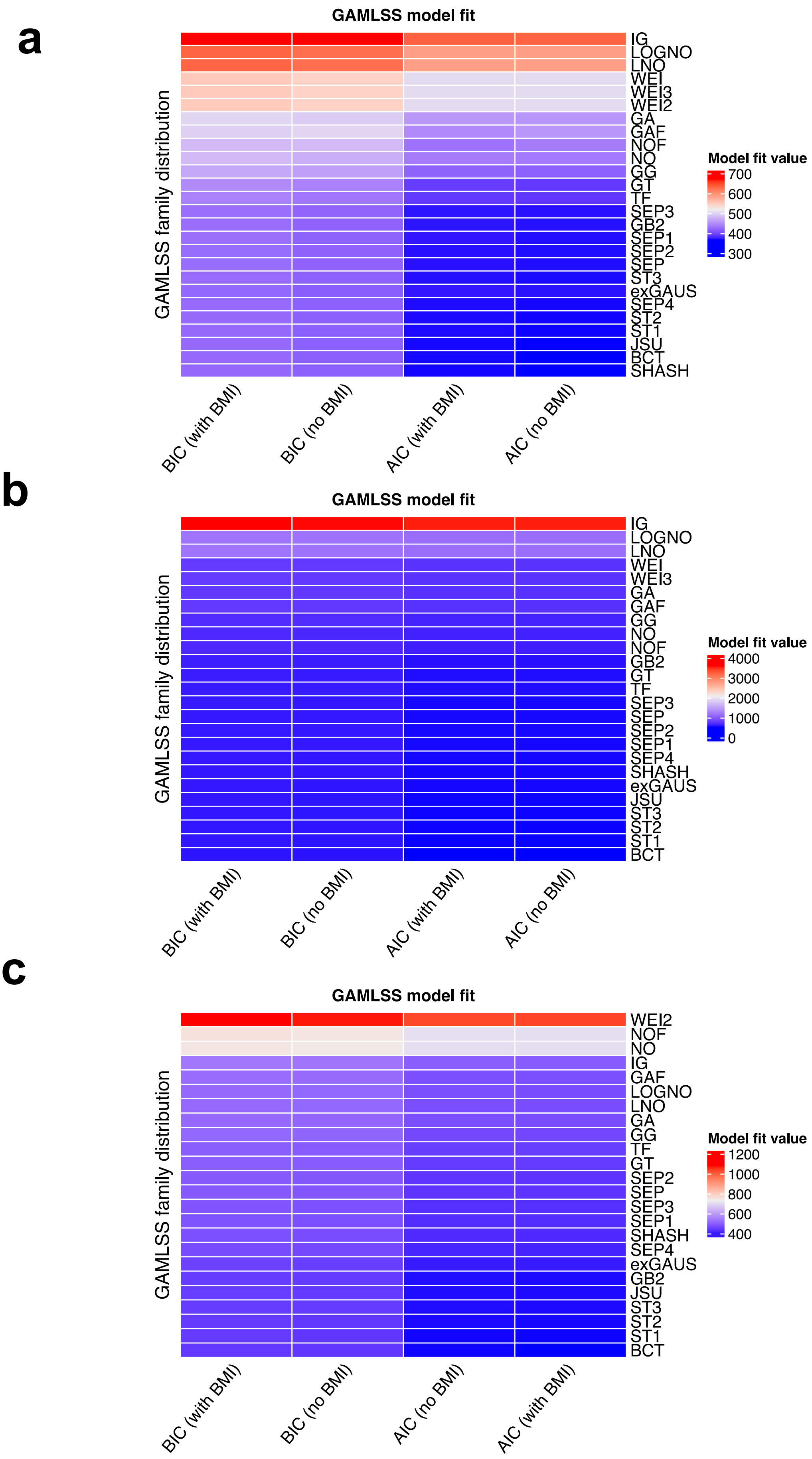

a
GAMLSS model fit
GAMLSS family distribution
IG
LOGNO
LNO
WEI
WEI3
WEI2
GA
GAF
NOF
NO
GG
GT
TF
SEP3
GB2
SEP1
SEP2
SEP
ST3
exGAUS
SEP4
ST2
ST1
JSU
BCT
SHASH
BIC (with BMI)
BIC (no BMI)
AIC (with BMI)
AIC (no BMI)
Model fit value
700
600
500
400
300
b
GAMLSS model fit
GAMLSS family distribution
IG
LOGNO
LNO
WEI
WEI3
GA
GAF
GG
NO
NOF
GB2
GT
TF
SEP3
SEP
SEP2
SEP1
SEP4
SHASH
exGAUS
JSU
ST3
ST2
ST1
BCT
BIC (with BMI)
BIC (no BMI)
AIC (with BMI)
AIC (no BMI)
Model fit value
4000
3000
2000
1000
0
c
GAMLSS model fit
GAMLSS family distribution
WEI2
NOF
NO
IG
GAF
LOGNO
LNO
GA
GG
TF
GT
SEP2
SEP
SEP3
SEP1
SHASH
SEP4
exGAUS
GB2
JSU
ST3
ST2
ST1
BCT
BIC (with BMI)
BIC (no BMI)
AIC (no BMI)
AIC (with BMI)
Model fit value
1200
1000
800
600
400

**Supplementary Figure 11. GAMLSS model fits for the effect of age on circulating retinol in the entire TwinsUK cohort**. Model fit parameters are denoted in the heatmap for retinol measurements from the first (**a**), second (**b**), and third (**c**) visits, respectively. The model fit parameters are the Bayesian information criterion (BIC) and Akaike information criterion (AIC), respectively, with models both with and without BMI added as an additional term in the model. Fit values are provided for a series of GAMLSS families: IG = inverse Gaussian, LOGNO = log-Normal, LNO = log-normal (Box-Cox), WEI = Weibull, WEI2 = Weibull (PH parameterisation), WEI3 = Weibull (mu as mean), GA = gamma, NO = Normal, GG = Generalised Gamma, GT = Generalised *t*, TF = *t*-distribution, SEP1 = Skew Power Exponential Type 1, SEP2 = Skew Power Exponential Type 2, SEP3 = Skew Power Exponential Type 3, SEP4 = Skew Power Exponential Type 4, ST1 = Skew *t* type 1, ST3 = Skew *t* type 2, ST3 = Skew *t* type 3, exGAUS = Exponential Gaussian, JSU = Johnson's SU, BCT = Box-Cox *t*, and SHASH = Sinh-Arcsinh.

a

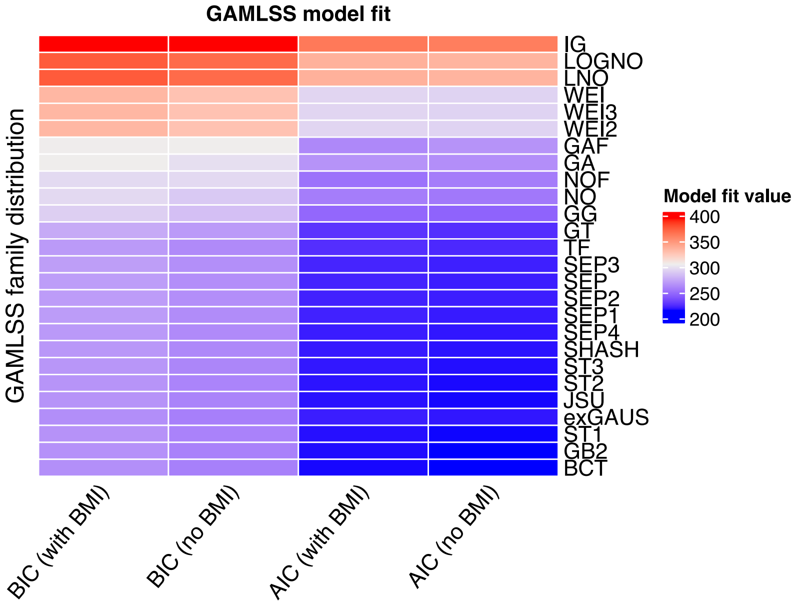
GAMLSS model fit
GAMLSS family distribution
IG
LOGNO
LNO
WEI
WEI3
WEI2
GAF
GA
NOF
NO
GG
GT
TF
SEP3
SEP
SEP2
SEP1
SEP4
SHASH
ST3
ST2
JSU
exGAUS
ST1
GB2
BCT
BIC (with BMI)
BIC (no BMI)
AIC (with BMI)
AIC (no BMI)
Model fit value
400
350
300
250
200

b

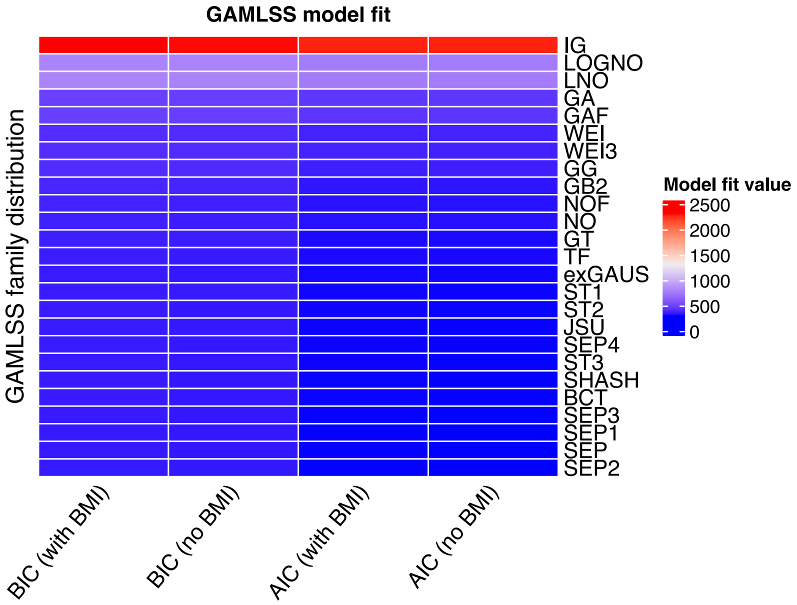
GAMLSS model fit
GAMLSS family distribution
IG
LOGNO
LNO
GA
GAF
WEI
WEI3
GG
GB2
NOF
NO
GT
TF
exGAUS
ST1
ST2
JSU
SEP4
ST3
SHASH
BCT
SEP3
SEP1
SEP
SEP2
BIC (with BMI)
BIC (no BMI)
AIC (with BMI)
AIC (no BMI)
Model fit value
2500
2000
1500
1000
500
0

c

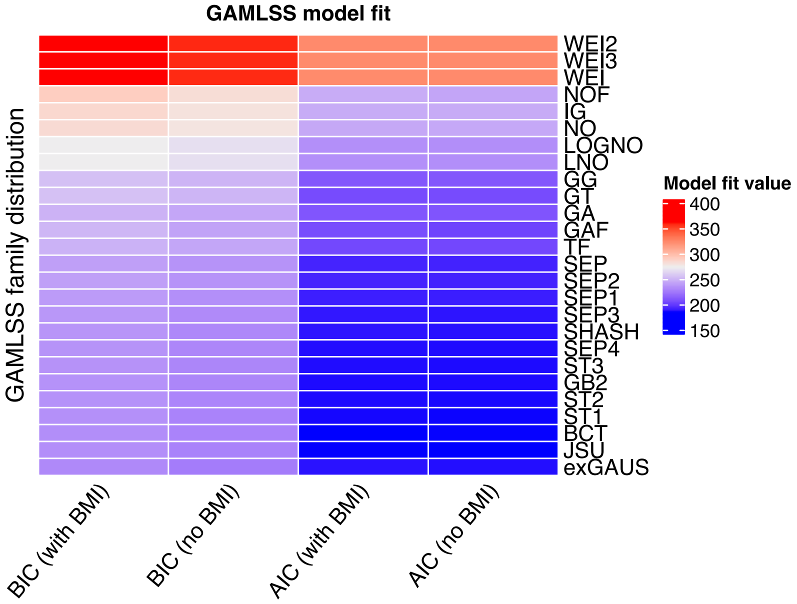
GAMLSS model fit
GAMLSS family distribution
WEI2
WEI3
WEI
NOF
IG
NO
LOGNO
LNO
GG
GT
GA
GAF
TF
SEP
SEP2
SEP1
SEP3
SHASH
SEP4
ST3
GB2
ST2
ST1
BCT
JSU
exGAUS
BIC (with BMI)
BIC (no BMI)
AIC (with BMI)
AIC (no BMI)
Model fit value
400
350
300
250
200
150

**Supplementary Figure 12. GAMLSS model fits for the effect of age on circulating retinol in half of TwinsUK cohort (unrelated individuals)**. Model fit parameters are denoted in the heatmap for retinol measurements from the first (**a**), second (**b**), and third (**c**) visits, respectively. The model fit parameters are the Bayesian information criterion (BIC) and Akaike information criterion (AIC), respectively, with models both with and without BMI added as an additional term in the model. Fit values are provided for a series of GAMLSS families: IG = inverse Gaussian, LOGNO = log-Normal, LNO = log-normal (Box-Cox), WEI = Weibull, WEI2 = Weibull (PH parameterisation), WEI3 = Weibull (mu as mean), GA = gamma, NO = Normal, GG = Generalised Gamma, GT = Generalised *t*, TF = *t*-distribution, SEP1 = Skew Power Exponential Type 1, SEP2 = Skew Power Exponential Type 2, SEP3 = Skew Power Exponential Type 3, SEP4 = Skew Power Exponential Type 4, ST1 = Skew *t* type 1, ST3 = Skew *t* type 2, ST3 = Skew *t* type 3, exGAUS = Exponential Gaussian, JSU = Johnson's SU, BCT = Box-Cox *t*, and SHASH = Sinh-Arcsinh.

### SUPPLEMENTARY TEXT

#### Gene prioritisation within the circulating retinol genome-wide significant loci

Full details of these results can be found in supplementary table 7.

***Locus 2:27598097-27752871***

In this locus on chromosome 2, most lines of evidence suggested that the known metabolic gene *GCKR* is associated with circulating retinol. The following lines of evidence were supportive of this inference:

- *GCKR* is the closest transcription start site (TSS) and the closest gene to the lead SNP.
- A non-synonymous variant exists with *GCKR*, in this case, it is the lead SNP. This SNP has been demonstrated previously to be functionally relevant for the GCKR protein.
- *GCKR* had the strongest finemapped pQTL of all variants in the locus.
- *GCKR* was supported by CADD score, RegulomeDB, and the V2G annotation pipeline.
- *GCKR* variants were exclusively in the 95% credible set with *PP* > 0.2 as derived from probabilistic finemapping.

The only evidence to support another gene in this locus was that the strongest (statistical significance) eQTL in the locus is associated with expression of the gene *KRTCAP3*. However, given the lack of other evidence, this is most likely driven by linkage, particularly as statistical significance does not imply causality in QTL relationships.

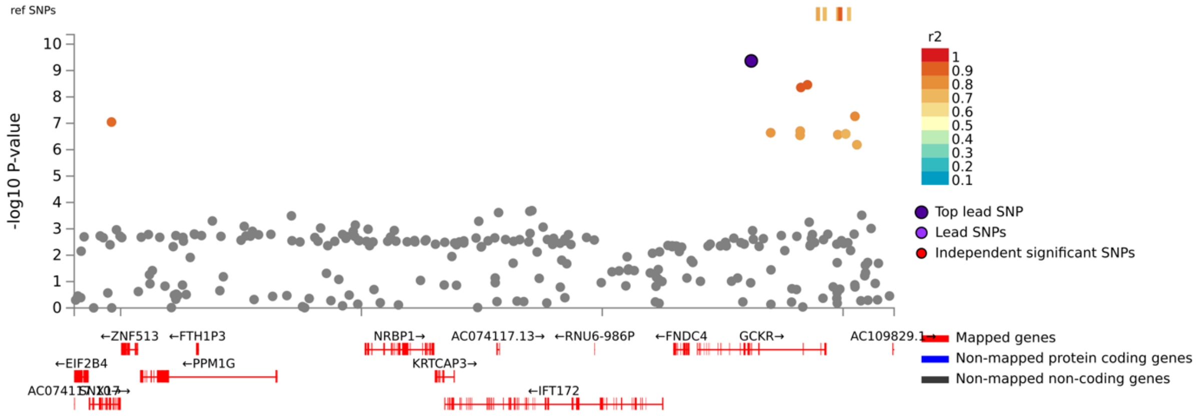

***Locus 2:122078406:122084285***

The gene prioritisation results within this locus were not conclusive. The closest TSS to the lead SNP was mapped to gene *TFCP2L1*, which has known significance for metabolic traits, and the most statistically significant eQTL in the locus also related to this gene. Despite this, many other lines of evidence like finemapping and the *in silico* annotation methods prioritised intergenic variants, suggesting further functional interrogation of this locus is warranted.

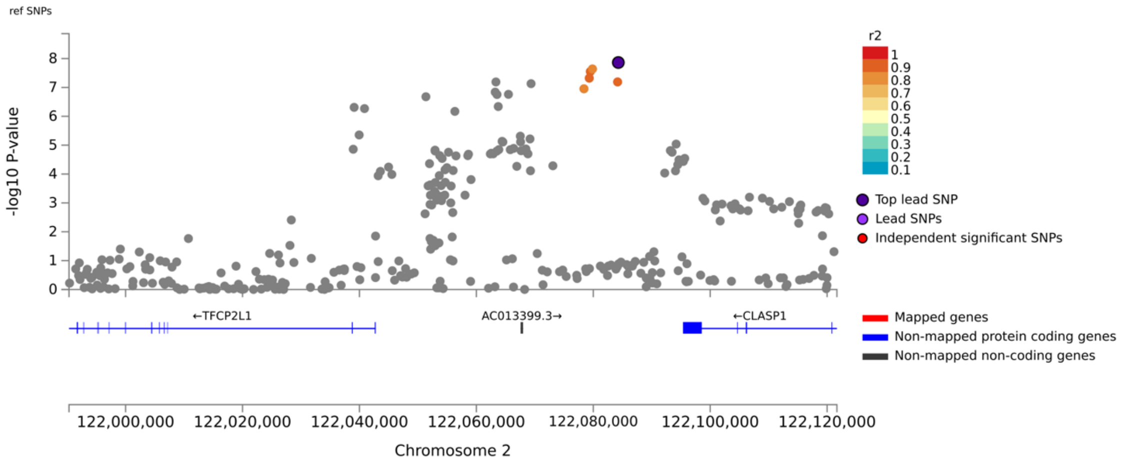


***Locus 7:114014488:114286611***

The transcription factor gene *FOXP2* had the strongest evidence within this locus, with no other genes confidently implicated. Specifically, it was supported by the following:

- Closest TSS and gene to lead SNP
- Most significant eQTL in locus associated with *FOXP2* expression
- Prioritised by CADD score, RegulomeDB, and the V2G pipeline
- Variants annotated to *FOXP2* in the 95% credible set.

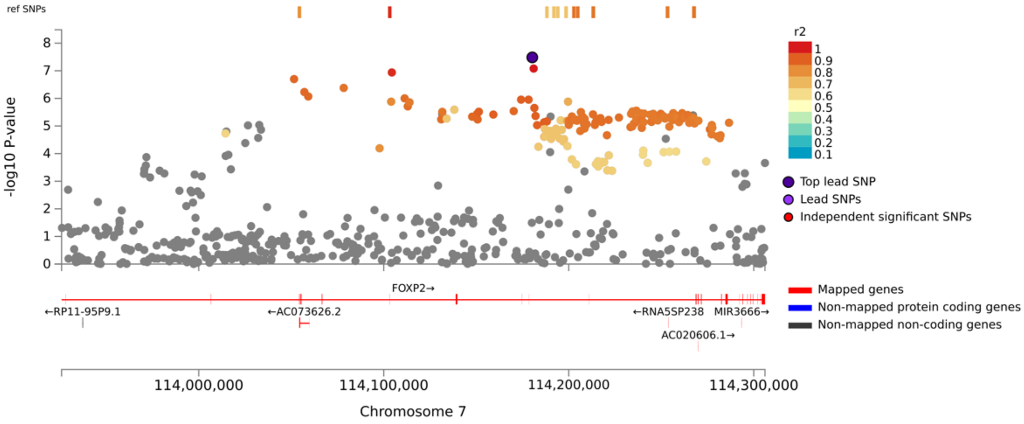

#### *Locus 8:9167797:9224907*

This locus was also not particularly conclusive as to a single gene that could be confidently prioritised. *PPP1R3B* was the closest TSS to the lead SNP, with variants in the 95% credible set annotated to a divergent non-coding transcript of this gene (*PPP1R3B-DT*). However, long non-coding RNA (lncRNA) like *RP11-115J16.1* were also supported by annotations like CADD score and RegulomeDB, with the locus peak overlapping a cluster of lncRNAs. As a result, this locus warrants future functional dissection to resolve its mechanistic relationship with circulating retinol.

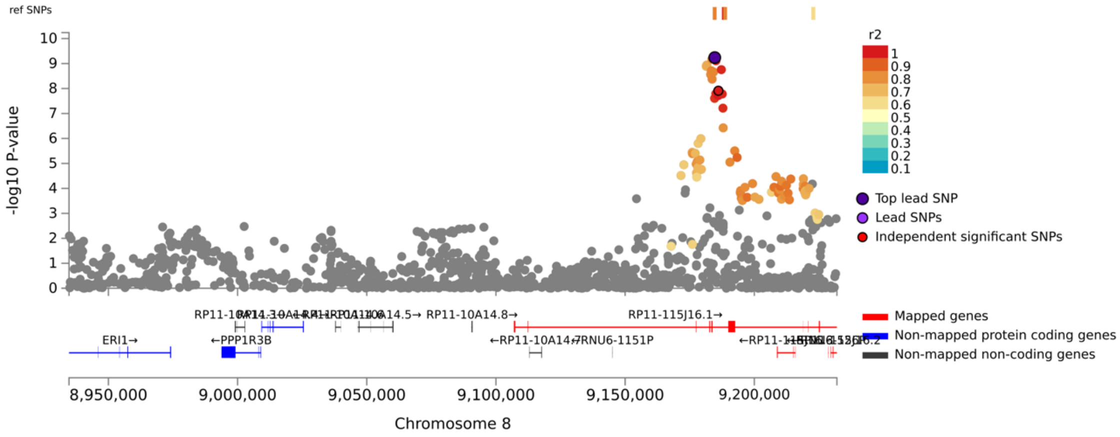


#### *Locus 10:95295876:95360964*

This association signal was one of the two previously implicated genome-wide significant signals associated with retinol. The causal gene at this locus is almost certainly *RBP4*, as would be expected given this gene encodes the principal serum retinol transporter. Whilst there is some evidence of eQTLs in the locus having a stronger statistical effect on the neighbouring gene *FFAR4*, the finemapped eQTLs and pQTLs map to *RBP4*. Additional effects on *FFAR4* should not be ruled given this is an important lipid gene and lipids are functionally related to retinol biology.

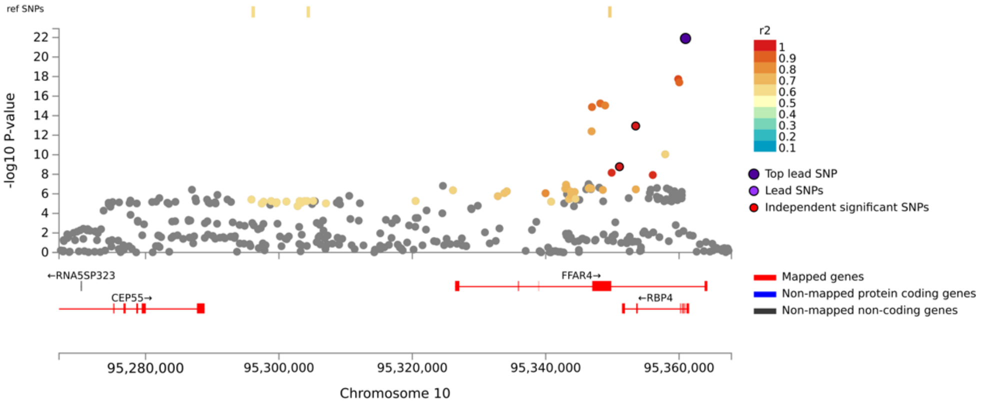

***Locus 16:79696939:79756197***

This locus had mixed evidence for different genes, however, as described in the main text, there is *in vitro* data to support the role of the *Maf* family of transcription factors given the genes implicated. A member of this family, *MAF*, is the closest TSS to the lead SNP. Two genes experimentally shown to regulate *MAF* (*MAFTRR* and *LINC01229*) are also implicated in this locus, with *MAFTRR* assigned a finemapped eQTL and *LINC01229* annotated to the 95% credible set.

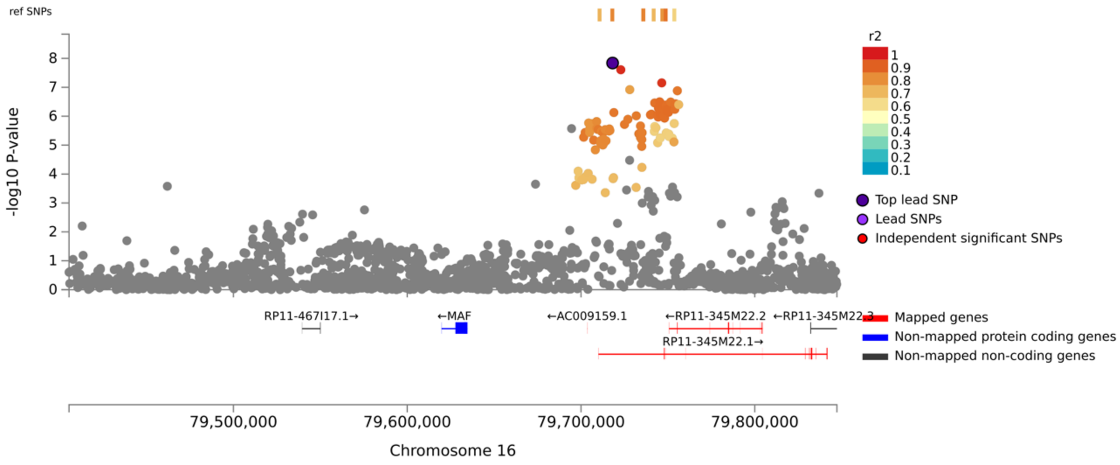


***Locus 18:29134171:29190174***

This is the second previously known retinol association signal and maps to *TTR*, which complexes with *RBP4* to form the *RBP4:TTR* complex that transports retinol in serum. There is some additional evidence in this locus to support other genes like *B4GALT6*, which could also be functionally relevant.

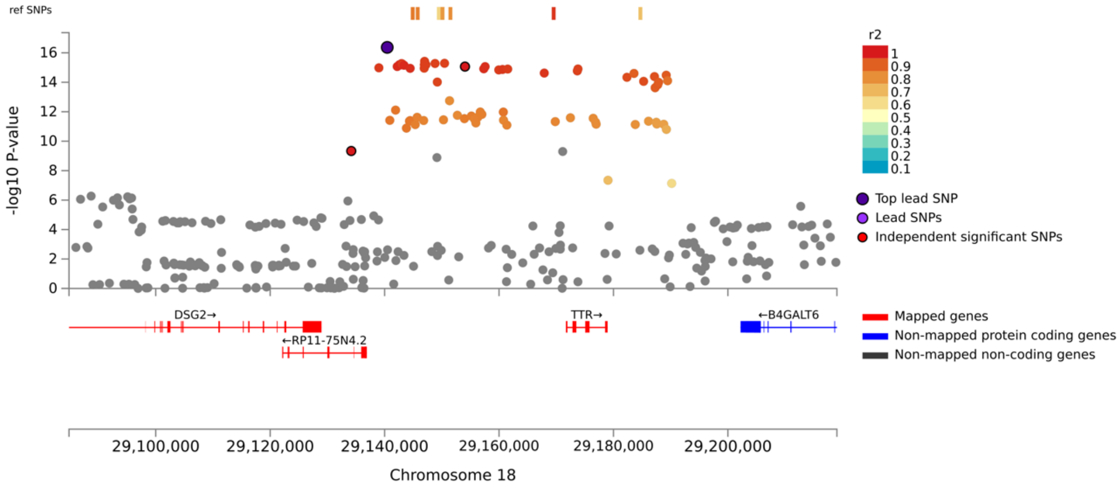

***Locus 20:39142516:39234223***

The only gene prioritised in this locus is another member of the *Maf* TF family (*MAFB*), which is the closest TSS to the lead SNP, the strongest eGene, and prioritised by V2G. However, several other lines of evidence support intergenic variants, and therefore, further investigation is needed before *MAFB* can be confidently assigned as a retinol associated gene.

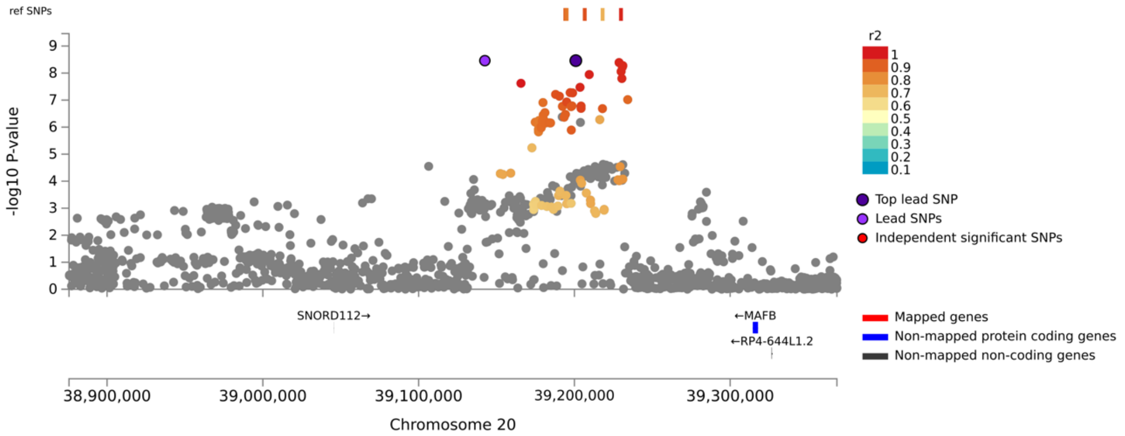


**Multivariable Mendelian randomisation models testing the effects of serum creatinine and major lipid species on circulating retinol**

We constructed multivariable Mendelian randomisation (MVMR) models to test the effect of serum creatinine on retinol conditioned on major lipid species (LDL, HDL, and triglycerides, main text figure 4B). After selecting independent variants associated with at least one of these traits at genome-wide significance, we calculated a conditional $F$ statistic for instrument strength. We were able to identify suitably powered instruments, although the conditional $F$ statistic for creatinine was just the suggested threshold of 10 after rounding to 1 decimal place ($F = 9.7$). As a result, the creatinine effects may be less reliable than the lipids, although given the univariable analyses strongly supported creatinine, we continued with this model. An inverse-variance weighted (IVW) MVMR model was first constructed. Triglycerides were found to have a non-zero effect on circulating retinol conditioned on the remaining exposures, estimated as a 0.153 [95% CI: 0.024, 0.28] SD increase in retinol per SD increase in circulating triglycerides. There was also a trend for a non-zero conditional effect of creatinine on retinol, but this was not quite statistically significant. Similar results were found using a multivariable extension of the MR Egger approach (mvEgger). However, the median (mvMedian) and penalised regression IV selection (mvLASSO) models instead supported that creatinine has a non-zero retinol increasing effect conditioned on lipids, whilst triglycerides only trended towards statistical significance. Therefore, we conclude that there is evidence to support an independent effect of both triglycerides and creatinine. In future, these relationships should be subjected to further interrogation to unravel their interrelationship with greater granularity.

**Profiling of plasma retinol in the INTERVAL cohort**

Plasma metabolites were measured using the DiscoveryHD4® platform (Metabolon, Inc., Durham, USA). Four batches of samples were prepared through random sampling from the INTERVAL study and metabolites were measured within these batches separately. The data was further 'jointly recalled' to update the metabolites to the current reference metabolomics library. Phenotype QC was performed within each batch. In the first step samples with missing values for each in 'OrigScale' were identified. These sample specific metabolite values were set to missing within the 'ScaledImpData' which contains for each metabolite the values within the 'OrigScale' median normalised for run day (median set to 1 for run-day batch). Metabolites were then excluded if measured in only one batch or in less than 100 samples. Metabolite values were then winsorized to 5 standard deviations from the mean where the values exceeded mean +/- 5 x standard deviation of the metabolite. Each metabolite was then log (natural) transformed prior to calculating the residuals adjusted for age, gender, INTERVAL centre, appointment month, the lag time between the blood donation appointment and sample processing and metabolon instrument batch (BATCH_400, BATCH_402, BATCH_209, and BATCH_305 columns per libraries 400, 402, 209, and 305). These residuals were standardised to a mean of 0 and standard deviation of 1 and rank normalised (rankNorm() function in R). Batch specific rank normalised values were then merged to create WGS and WES specific phenotype files prior to the genetic analyses performed in HAIL adjusted for INTERVAL metabolon batch and 10 genetic PCs.

**Comparing the inverse variance weighted estimator with multiplicative random effects versus fixed effects in the absence of instrumental variable heterogeneity**

We observe residual standard errors < 1 in IVW-MRE models that are Tier #2 or Tier # 3, and therefore, do not exhibit statistically non-zero heterogeneity between IV exposure-outcome estimates. This is reflected by the IVW fixed effects for standard errors for these trait pairings being larger than that of the IVW-MRE, although the magnitude of this does vary, with some traits exhibiting a larger difference between the two methods than others. In the MR literature, residual standard error < 1 is characterised as 'under-dispersion', and what gives rise to this is not immediately clear. However, it may be the case for metabolite traits like retinol, where the biology is more defined relative to more complex traits, it is easier to define instrumental variables that have more consistent exposure-outcome relationships. It could also be a chance finding. Regardless, we chose to further investigate the implications for our multiple-testing correction upon using a fixed effects IVW model for those exposure-outcome pairs which do not exhibit non-zero heterogeneity. This was achieved by testing for heterogeneity via Cochran's Q before the IVW and using a fixed effects estimator for traits without significant heterogeneity and MRE for those that do exhibit heterogeneity. FDR correction was then applied across the combined results of both tests such that each trait had a single IVW *P*-value (fixed effects or MRE). Several of the tier #2/tier #3 traits had higher fixed effects *P*-values, and therefore, a higher FDR in these analyses. However, overall, very similar biology was prioritised (FDR < 0.1) – specifically body/trunk-fat percentage, keratometry measurements, the microbiome, MRI phenotypes, and arthrosis of the hip.

**Normative modelling**

***Selection of the GAMLSS family to construct the normative models***

We used a pragmatic approach to select the optimal GAMLSS distribution family through calculating both the AIC and BIC (Supplementary Figures 9 and 10). There were only very nominal differences in overall model fit between several of the tested families. We chose the Box-cox *t* distribution GAMLSS family as it performed best over the majority of tested visits in the full cohort, as well as showing good performance in half of the twins. We also found that adding BMI as an additional term did not greatly improve model fit, and thus, only age and batch were modelled in the subsequent steps of our framework.
